# Assessing the role of local health departments in implementing PrEP in the US, 2020 update

**DOI:** 10.1101/2024.05.23.24307795

**Authors:** Julia R. Zigman, Karen W. Hoover, Rupa R. Patel, Kellie Hall, Roman J. Gvetadze, Ya-lin A. Huang

## Abstract

Objectives

To identify current and potential roles for, and systematic barriers encountered by, local health departments (LHDs) engaging in PrEP implementation.

**Methods:** Web-based assessment created with Qualtrics® from Oct.-Dec. 2020 distributed via e-mail to 1,096 LHDs that indicated they provided HIV or STD screening and/or treatment in NACCHO’s 2019 National Profile of Local Health Departments.

**Results:** Of 354 respondents, 46% of LHDs were engaged in PrEP implementation compared with 29% in 2015. Activities included: referring to PrEP providers (68%); conducting consumer education (65%); promoting PrEP (59%); educating providers; (48%); and prescribing PrEP (42%). The top challenges to initiating or expanding PrEP implementation were lack of funding and staff, and 41% described staff reductions during COVID-19.

**Conclusions:** LHD engagement in PrEP implementation has increased significantly since 2015 but many LHDs still encounter barriers to initiating or expanding PrEP activities.

**Policy Implications:** Robust funding of public health infrastructure can ensure that LHDs and their partners are equipped to provide PrEP access for all who may benefit from it during future public health emergencies.

## Introduction

Preexposure prophylaxis (PrEP) is a biomedical intervention for the prevention of HIV. The initial PrEP medication approved by the Food and Drug Administration in 2012, a daily oral pill of two co-formulated antiretroviral drugs, was recommended for men and women who are at risk for HIV, by the Centers for Disease Control and Prevention (CDC) in 2014.(1) Since then, two additional agents for PrEP have been approved including an injectable long-acting antiretroviral medication.(1) The use of PrEP has increased markedly in the United States since it became available, with 48% of persons with PrEP indications prescribed it in 2022.(2) But disparities in PrEP use by persons in racial and ethnic minority populations and women have persisted.(2) The public health community is responding to a call to action to reduce these disparities in order to achieve the Ending the HIV Epidemic in the U.S. (EHE) goal to decrease new HIV infections by 90% by 2030.(3)

State and local health departments play a critical role in HIV prevention in the United States including the implementation of PrEP. Supported by federal, state, and local governments and other entities, the health department conducts a multitude of HIV prevention services and activities.(4) These activities include, but are not limited to: direct and community-based organization (CBO) supported community outreach; sexually transmitted infections (STIs), PrEP, and other clinical services; HIV testing; linkage of persons in the community to PrEP clinical services; laboratory testing support for HIV, STI, hepatitis, and other conditions; provider and community education; partner services; and liaising with community groups.

Understanding the needs of health departments can help CDC support them to achieve both increased PrEP implementation and EHE goals for all populations. Increased understanding of health department needs can help to guide the development of funding announcements and CDC’s capacity building resources. In 2015, the National Association of County and City Health Officials (NACCHO) and CDC conducted a survey of U.S. local health departments (LHDs) to understand their activities to implement PrEP and support its use by persons in their jurisdictions.(5) Responses revealed that support of PrEP implementation was in early stages at most health departments, and specific resource needs were reported including materials to support healthcare providers to prescribe PrEP, materials for community education about HIV and PrEP, and protocols for PrEP referrals. We repeated a modified version of the 2015 survey in 2022 to assess the progress made by health departments in their PrEP implementation programs and to understand ongoing challenges and resource needs.

## Methods

NACCHO and CDC conducted an inaugural web-based survey of approximately 500 LHDs assessing their engagement in and support for PrEP implementation in 2015. We conducted a second iteration of this web-based survey in 2020 to examine LHD engagement in PrEP implementation before and during the COVID-19 pandemic. The NACCHO team developed the 2020 instrument by first reviewing the previous (i.e., 2015) survey to determine which questions to keep, which to modify, and whether any new questions should be added. We piloted the 2020 survey with nine LHDs recruited from NACCHO advisory groups and finalized the questions based on their feedback. The sampling method used in 2020 was similar to that of 2015. The survey sample was drawn from 1,463 LHDs which indicated that they provide or contract out HIV or STI screening, treatment or family planning services in the NACCHO’s 2019 National Profile of Local Health Departments (Profile) study.(6) LHDs receiving multiple NACCHO surveys were removed from the sample to reduce survey burden. The sampling frame was stratified by U.S. census region and population size served by the LHD (<50,000; 50,000– 499,999; 500,000+).(6) Between October and December 2020, we distributed the survey to 1,096 LHDs using Qualtrics. The survey was e-mailed to the LHD’s primary contact, or the individual designated to respond to inquiries about PrEP. After the initial e-mail invitation, each LHD representative received up to nine reminder emails and reminder calls. The primary objective of this assessment was to understand the mechanisms through which LHDs engage in PrEP implementation, and secondary objectives included assessing the impact of COVID-19 and other systemic challenges on the ability to implement these mechanisms, as well as identifying changes in the prevalence of, and mechanisms used for, LHD engagement in PrEP implementation from 2015 to 2020.

### Measures

The 2020 survey included questions assessing LHD HIV prevention program structure, services, and engagement in PrEP implementation. We defined LHDs that were supporting PrEP implementation as engaged. This broad definition of engagement in PrEP included any activities to support PrEP delivery (e.g., education and outreach, referral and linkage, prescribing, monitoring, planning). LHDs that indicated they were engaged in PrEP received a module questionnaire, assessing specific PrEP implementation activities, resources needed to advance PrEP implementation, and the impact of COVID-19 on PrEP-related services. LHDs that indicated that they were not engaged in PrEP implementation received an alternate module questionnaire, assessing their awareness and interest in PrEP, barriers to future PrEP engagement, resources needed to initiate PrEP implementation, and perceived potential roles in future PrEP engagement. Finally, all LHDs regardless of PrEP engagement received a third module questionnaire, assessing familiarity with patient assistance programs for PrEP and pre- and post-COVID-19 HIV testing policies and strategies.

The primary objective was measured by analyzing the prevalence of reported methods of, and challenges to, PrEP engagement by LHDs. The change in LHD engagement in PrEP was measured by comparing repeated and/or rephrased questions between the 2015 and 2020 versions of the assessment, while the impact of COVID-19 was assessed via specific descriptive questions.

### Data analysis

We reported descriptive statistics and weighted proportions for all LHDs responding to the survey, stratified by LHD’s PrEP engagement. We compared the weighted responses between the 2015 and 2020 surveys and tested the differences using Rao-Scott chi-square tests. Estimates from both years were weighted similarly to produce nationally representative estimates, accounting for the stratified survey design in each year. All the analyses were performed using Stata® version 15.1 (StataCorp) and SAS version 9.4 (Cary, NC).

This activity was reviewed by CDC, deemed not research, and was conducted consistent with applicable federal law and CDC policy.

## Results

A total of 353 LHDs across all U.S. Census regions responded to the 2020 survey, comprising a 32% response rate. Forty-three (43%) respondents represented LHDs serving small populations (<50,000), with 45% and 12% serving medium (50,000–499,999) and large (>500,000) populations respectively.

Engagement in PrEP implementation among surveyed LHDs increased from 29.2% in 2015 to 45.8% in 2020 (p < 0.0001). A greater proportion of LHDs engaged in PrEP implementation if they were in larger jurisdictions or operated STI clinics (**Table 1**).

**Table 1.** Characteristics of local health departments responding to PrEP implementation surveys and involvement in PrEP over survey years, United States, 2015 and 2020.

| Characteristic | 2015 NACCHO Survey: Involvement in PrEP |  |  | 2020 NACCHO Survey: Involvement in PrEP |  |  | Rao-Scott<br>ChiSq<br>p-value |
| --- | --- | --- | --- | --- | --- | --- | --- |
|  | LHD (n=284)<br>Unweighted<br>n (%) | Engaged (n=109)<br>Weighted %<br>(95% CI) | Not engaged (n=175)<br>Weighted %<br>(95% CI) | LHD (n=353)<br>Unweighted<br>n (%) | Engaged (n=171)<br>Weighted %<br>(95% CI) | Not engaged (n=182)<br>Weighted %<br>(95% CI) |  |
| All respondents | 284 (100.0) | 29.2 (23.3, 35.1) | 70.8 (64.9, 76.7) | 353 (100.0) | 45.8 (40.6, 51.0) | 54.2 (49.0, 59.4) | <.0001 |
| **Jurisdiction size** |  |  |  |  |  |  |  |
| Small | 98 (34.5) | 17.9 (10.0, 25.8) | 82.1 (74.2, 90.0) | 150 (42.5) | 36.0 (28.3, 43.7) | 64.0 (56.3, 71.7) | 0.0024 |
| Medium | 116 (40.8) | 32.0 (22.4, 41.7) | 68.0 (58.3, 77.6) | 160 (45.3) | 50.0 (42.2, 57.8) | 50.0 (42.2, 57.8) | 0.0055 |
| Large | 70 (24.6) | 68.0 (57.0, 79.1) | 32.0 (20.9, 43.0) | 43 (12.2) | 86.0 (75.7, 96.4) | 14.0 (3.6, 24.3) | 0.0325 |
| **Census region** |  |  |  |  |  |  |  |
| West | 60 (21.1) | 46.8 (33.9, 59.8) | 53.2 (40.2, 66.1) | 44 (12.5) | 58.0 (42.9, 73.0) | 42.0 (27.0, 57.1) | 0.2734 |
| South | 117 (41.2) | 33.3 (24.3, 42.3) | 66.7 (57.7, 75.7) | 175 (49.6) | 51.1 (43.6, 58.6) | 48.9 (41.4, 56.4) | 0.0040 |
| Northeast | 62 (21.8) | 36.7 (24.7, 48.7) | 63.3 (51.3, 75.3) | 32 (9.1) | 44.8 (27.4, 62.3) | 55.2 (37.7, 72.6) | 0.4448 |
| Midwest | 45 (15.8) | 15.9 (5.2, 26.5) | 84.1 (73.5, 94.8) | 102 (28.9) | 32.5 (23.5, 41.6) | 67.5 (58.4, 76.5) | 0.0356 |
| **Operates STI clinic** |  |  |  |  |  |  |  |
| Yes | 201 (70.8) | 36.5 (28.8, 44.2) | 63.5 (55.8, 71.2) | 157 (44.5) | 65.3 (57.7, 73.0) | 34.7 (27.0, 42.3) | <.0001 |
| No | 81 (28.5) | 15.9 (8.0, 23.7) | 84.1 (76.3, 92.0) | 133 (37.7) | 35.6 (27.3, 43.8) | 64.4 (56.2, 72.7) | 0.0011 |

LHD: Local Health Department; CI: Confidence Interval; PrEP: Preexposure prophylaxis; NACCHO: National Association of County and City Health Officials 2015/2020 Survey
Notes: The unweighted values do not always add to total because of missing responses or rounding error. The weighted percentage in each cell is computed using the total number of non-missing responses in the denominator

### Engagement in PrEP implementation

Respondents were asked to identify specific activities related to PrEP implementation engaged in by their LHDs. Frequently reported activities included referring or linking individuals to PrEP providers (69%), conducting consumer education and outreach (65%), using health communication methods and strategies to promote PrEP (59%), and providing education and/or training to healthcare providers (48%). Notably, the proportion of LHDs prescribing PrEP in a health department clinic more than quadrupled between 2015 (9%) and 2020 (42%) (p < 0.0001). Less frequently reported activities included maintaining or actively updating a directory of local PrEP providers (42%); providing PrEP adherence support (32%), providing routine HIV/STI screening for patients prescribed PrEP by a non-health department provider (32%), monitoring PrEP uptake (17%), and funding community partners to provide PrEP services (5%) (**Table 2**).

**Table 2.** Local health department PrEP implementation activities and optimal roles, United States, 2015 and 2020, weighted response proportions.

| Implementation Activities and Roles | Activities |  |  | Optimal Roles |  |  |
| --- | --- | --- | --- | --- | --- | --- |
|  | Engaged in PrEP in 2015 (n=109)<br>Weighted %<br>(95% CI) | Engaged in PrEP in 2020 (n=171)<br>Weighted %<br>(95% CI) | Rao-Scott<br>ChiSq<br>p-value | Engaged in PrEP in 2015 (n=109)<br>Weighted %<br>(95% CI) | Engaged in PrEP in 2020 (n=171)<br>Weighted %<br>(95% CI) | Rao-Scott<br>ChiSq<br>p-value |
| Conduct consumer education and outreach | 51.4 (40.0, 62.8) | 65.1 (57.5, 72.6) | 0.0513 | 58.2 (46.8, 69.5) | 59.1 (51.5, 66.8) | 0.8885 |
| Provide education and/or training to healthcare providers | 40.0 (28.8, 51.1) | 48.0 (40.3, 55.8) | 0.2479 | 54.2 (42.8, 65.7) | 43.8 (36.2, 51.5) | 0.1349 |
| Maintain or actively update a directory of local PrEP providers | 44.6 (33.4, 55.7) | 41.7 (34.1, 49.3) | 0.6769 | 59.1 (48.2, 69.9) | 44.5 (36.8, 52.1) | 0.0328 |
| Refer or link individuals to PrEP providers | 74.4 (64.5, 84.2) | 68.5 (61.1, 75.9) | 0.3561 | 75.9 (66.1, 85.6) | 59.0 (51.4, 66.6) | 0.0115 |
| Prescribe PrEP via health department clinic | 9.0 (3.5, 14.5) | 41.8 (34.3, 49.4) | <.0001 | 27.1 (18.2, 36.1) | 46.4 (38.9, 54.0) | 0.0016 |
| Fund community partners to provide PrEP services | 3.0 (0.5, 5.4) | 5.1 (2.2, 8.1) | 0.2790 | 6.1 (2.5, 9.8) | 7.0 (3.5, 10.4) | 0.7531 |
| Monitor PrEP uptake | 8.9 (3.7, 14.1) | 16.8 (11.2, 22.4) | 0.0512 | 23.3 (15.2, 31.4) | 23.5 (17.0, 30.0) | 0.9714 |
| Other optimal roles | 5.9 (2.0, 9.7) | 2.8 (0.2, 5.3) | 0.1900 | 1.1 (0.0, 2.6) | 5.8 (2.3, 9.3) | 0.0113 |
| Our optimal role is what we are currently doing to support PrEP implementation | *Not asked* | *Not asked* |  | 15.6 (7.2, 24.0) | 44.1 (36.5, 51.6) | <.0001 |
| Use health communication methods and strategies to promote PrEP | *Not asked* | 59.3 (51.8, 66.9) |  | *Not asked* | 66.3 (58.8, 73.7) |  |
| Provide PrEP adherence support | *Not asked* | 32.0 (24.9, 39.0) |  | *Not asked* | 43.5 (36.4, 50.6) |  |
| Provide routine HIV/STI screening for patients prescribed PrEP by a non-health department provider | *Not asked* | 32.1 (25.0, 39.3) |  | *Not asked* | 44.3 (36.7, 51.9) |  |
| Internal training for health department staff | 35.8 (25.0, 46.5) | *Not asked* |  | 39.9 (28.5, 51.4) | *Not asked* |  |
| Convene or participate in a working group on PrEP | 32.4 (22.1, 42.7) | *Not asked* |  | 30.8 (20.9, 40.6) | *Not asked* |  |
| Collaborate with health care providers to support PrEP delivery | 44.7 (33.3, 56.1) | *Not asked* |  | 54.8 (43.1, 66.5) | *Not asked* |  |
| Participation in a demonstration project or implementation study | 4.7 (1.5, 7.8) | *Not asked* |  | 13.6 (7.6, 19.6) | *Not asked* |  |

PrEP: Preexposure prophylaxis, STI: Sexually Transmitted Infection

Respondents also reported what they perceived to be their LHD’s optimal roles in PrEP implementation. Engaged LHDs in 2020 most frequently reported their optimal role as conducting PrEP promotion (66%), where in 2015 their most optimal role was referring or linking individuals to PrEP (76%), which is still frequently reported as an optimal role in 2020 but at a lower level (59%, p < 0.01). The proportion of engaged LHDs who reported in 2020 that their optimal role was prescribing PrEP at the health department clinic (46%) almost doubled since 2015 (27%) (p < 0.002). In 2020, 44% of engaged LHDs reported that they consider their current PrEP implementation activities to be an optimal role compared with 16% in 2015.

### Barriers to PrEP Engagement

Both engaged and non-engaged LHDs were asked about barriers to expanding or initiating engagement in PrEP implementation. In 2020, a lack of funding was the most reported challenge by both engaged (56%) and non-engaged (42%) LHDs (**Table 3**). Similarly, 57% of engaged LHDs listed lack of staff as a challenge to PrEP implementation. Other major challenges for engaged LHDs included difficulty reaching underserved populations for PrEP (45%) and lack of healthcare providers in the community prescribing PrEP (52%). Among non-engaged LHDs, significant challenges to initiating PrEP engagement included lack of a plan or strategy for PrEP implementation (39%), concerns related to reimbursement for PrEP clinical services (37%), and client access to financial resources to pay for PrEP (33%). (**Table 3**).

**Table 3.**
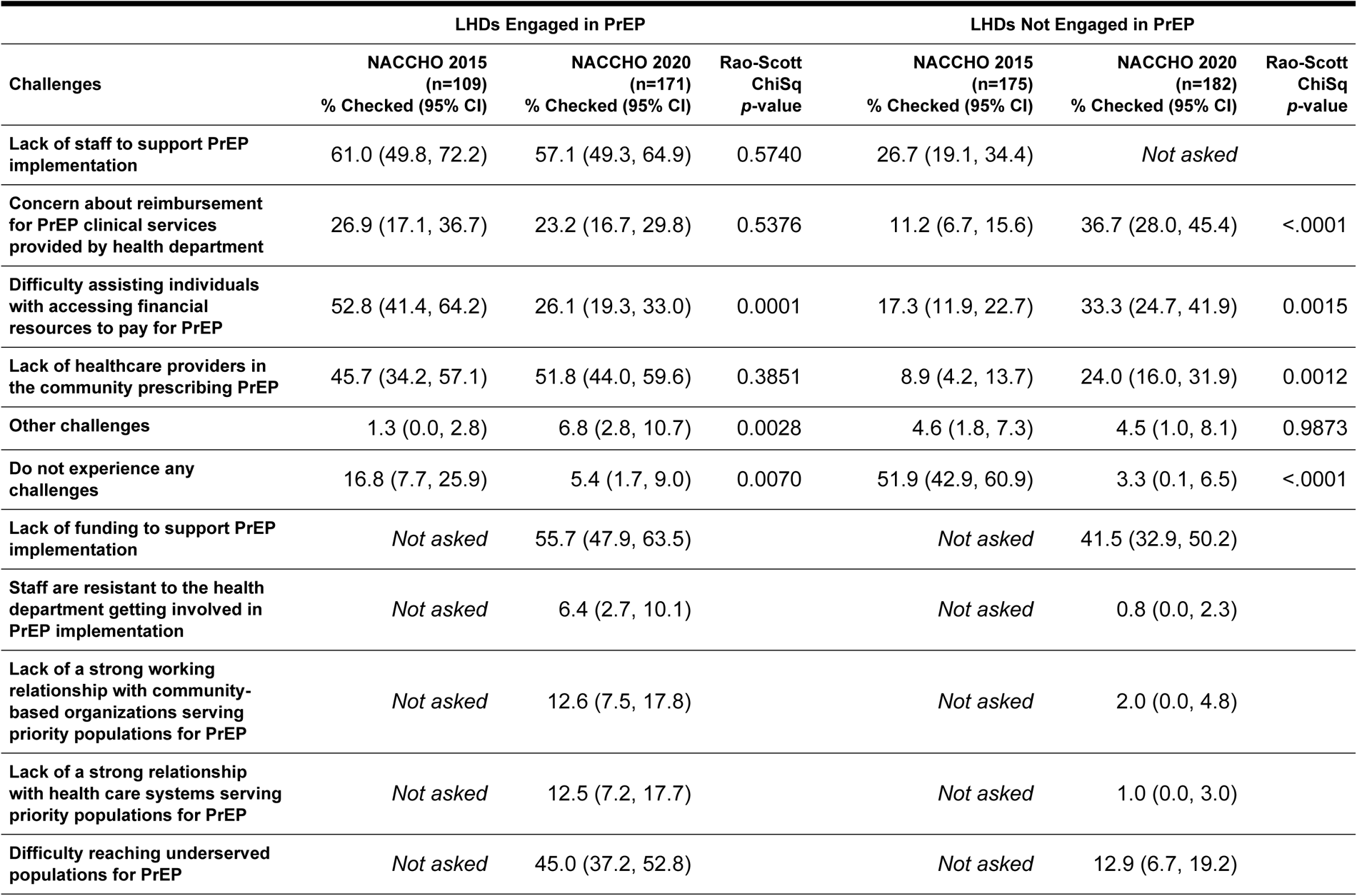
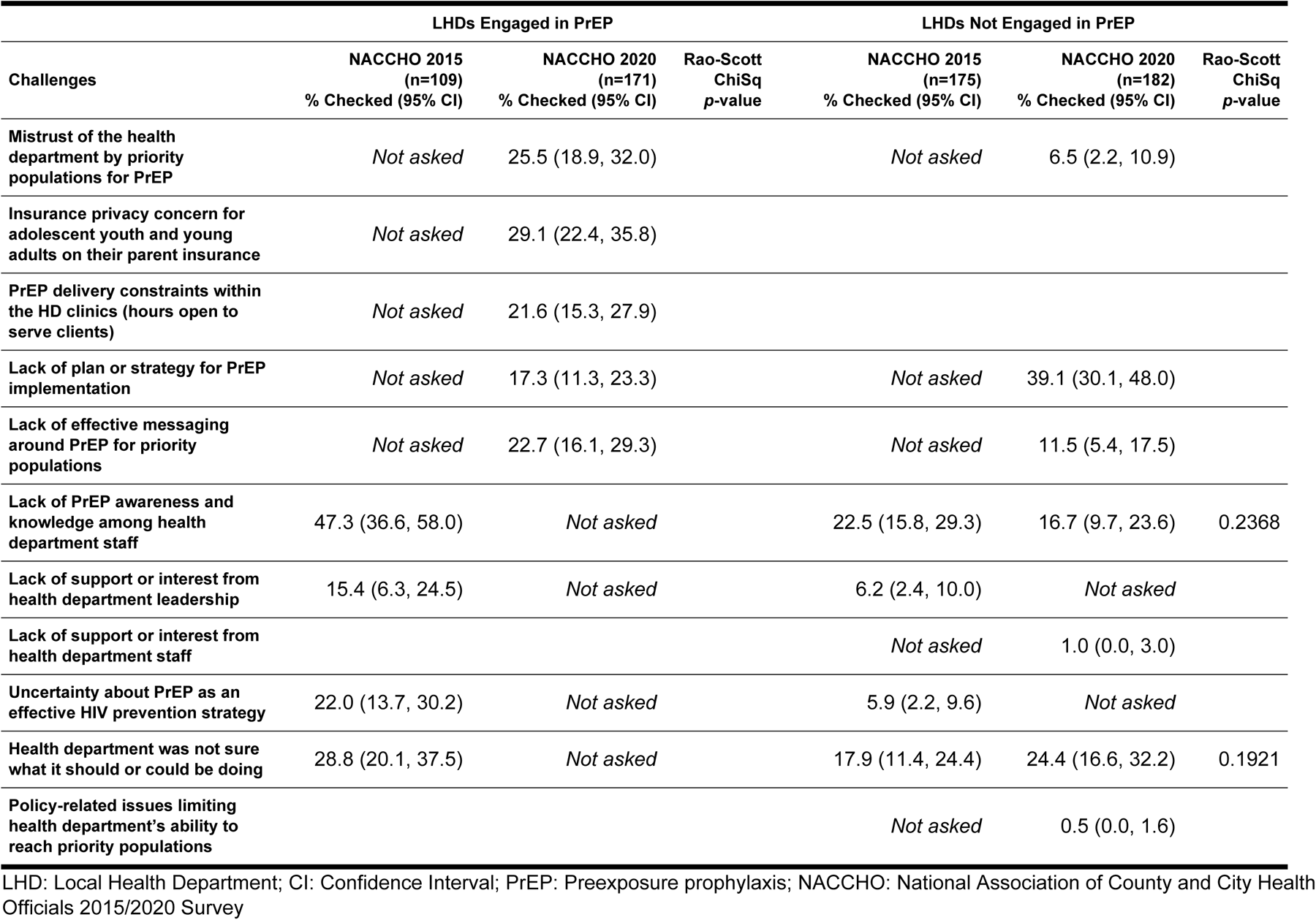
Challenges to PrEP implementation for local health departments, United States, 2015 and 2020, weighted response proportions, year-to-year comparison.

### Funding and staffing

Almost one-third (32%) of LHDs reported that that there was no specific funding to support their PrEP activities. Moreover, fewer than half (40%) of LHDs engaged in PrEP implementation had a staff position of at least 0.5 fulltime employees that was responsible for coordinating PrEP activities, with large jurisdictions being four times (80%) more likely to have this staff capacity compared with small jurisdictions (17%) (p<0.0001).

Non-engaged LHDs broadly reported increased challenges in 2020 versus 2015: in 2015, 52% of non-engaged LHDs reported they didn’t face any challenges to engaging in PrEP implementation compared with 3% in 2020 (p<0.0001). The proportion of non-engaged LHDs that reported concerns about reimbursement tripled from 2015 (11%) to 2020 (37%) (p<0.0001); similarly, twice as many non-engaged LHDs reported challenges assisting individuals with accessing financial resources to pay for PrEP from 2015 (17%) to 2020 (33%) (p=0.0015). However, it’s notable that among *engaged* LHDs, significantly fewer reported this challenge in 2020 (26%) than in 2015 (53%) (p<0001). Lastly, almost a quarter of non-engaged LHDs report that the health department is not sure what it should or could be doing (24%) (**Table 3**).

### Future funding uses

Both engaged and non-engaged LHDs were asked what they would prioritize if funding became available to support PrEP implementation. Echoing the 2015 survey, LHDs that had not yet engaged in PrEP implementation in 2020 most frequently reported that they would prioritize funding for PrEP planning (67%), followed by education and outreach to priority populations for PrEP (50%), and healthcare provider education (45%). Besides planning, these responses were similar among LHDs already engaged in PrEP implementation in 2020, among which education and outreach to priority populations (70%) and healthcare provider education (47%) were the most reported priorities, followed by providing PrEP adherence support (38%) and hiring clinical personnel (35%). The proportion of engaged LHD that reported prioritizing funding for education and outreach more than doubled from 2015 (31%) to 2020 (70%) (p=0.0003). (**Table 4**).

**Table 4.** Priority areas for funding PrEP implementation by local health departments, United States, 2015 and 2020, weighted response Proportions.

| Priority area for funding | LHDs Engaged in PrEP |  |  | LHDs Not Engaged in PrEP |  |  |
| --- | --- | --- | --- | --- | --- | --- |
|  | NACCHO 2015<br>(n=109)<br>% Checked (95% CI) | NACCHO 2020<br>(n=171)<br>% Checked (95% CI) | Rao-Scott<br>ChiSq<br>p-value | NACCHO 2015<br>(n=175)<br>% Checked (95% CI) | NACCHO 2020<br>(n=182)<br>% Checked (95% CI) | Rao-Scott<br>ChiSq<br>p-value |
| Plan for how to most effectively incorporate PrEP into HIV prevention education and services | 42.3 (28.0, 56.7) | *Not asked* |  | 68.9 (55.5, 82.4) | 66.7 (57.9, 75.6) | 0.7907 |
| <sup>1</sup> Education and outreach to priority populations for PrEP | 31.4 (14.7, 48.1) | 70.1 (62.8, 77.4) | 0.0003 | 36.2 (22.8, 49.5) | 50.3 (40.8, 59.8) | 0.0968 |
| Communications activities related to PrEP | 31.9 (18.1, 45.7) | *Not asked* |  | 22.1 (11.3, 32.8) | *Not asked* |  |
| Healthcare provider education | 60.7 (45.6, 75.9) | 46.8 (38.8, 54.8) | 0.1106 | 43.5 (29.6, 57.4) | 45.4 (36.3, 54.5) | 0.8155 |
| <sup>2</sup> Provide PrEP adherence support | 25.4 (12.1, 38.6) | 37.9 (30.2, 45.6) | 0.1344 | 29.9 (16.9, 42.8) | 24.8 (16.8, 32.8) | 0.5004 |
| <sup>3</sup> Monitoring PrEP uptake | 45.1 (29.6, 60.6) | 17.8 (11.8, 23.9) | 0.0009 | 16.8 (6.3, 27.3) | 13.1 (6.8, 19.3) | 0.5195 |
| Program staff to conduct non-clinical PrEP-related activities (e.g., assessment of eligibility for insurance, navigation or linkage to providers) | 46.7 (32.6, 60.8) | *Not asked* |  | 28.8 (15.8, 41.7) | *Not asked* |  |
| Provide HIV, and STI screening for patients prescribed PrEP by a non-health department provider | *Not asked* | 16.1 (10.1, 22.1) |  | *Not asked* | 35.3 (26.5, 44.1) |  |
| Hire clinical personnel | 28.4 (16.2, 40.5) | 34.9 (27.6, 42.2) | 0.3674 | 32.8 (19.8, 45.8) | 25.1 (17.1, 33.2) | 0.3116 |
| Hire non-clinical personnel | *Not asked* | 19.6 (13.6, 25.7) |  | *Not asked* | 8.4 (3.1, 13.7) |  |
| Provide funding to community partners to support PrEP implementation | *Not asked* | 18.0 (12.1, 23.8) |  | *Not asked* | 16.7 (9.8, 23.6) |  |
|  | NACCHO 2015<br>(n=109)<br>% Checked (95% CI) | NACCHO 2020<br>(n=171)<br>% Checked (95% CI) | Rao-Scott<br>ChiSq<br>p-value | NACCHO 2015<br>(n=175)<br>% Checked (95% CI) | NACCHO 2020<br>(n=182)<br>% Checked (95% CI) | Rao-Scott<br>ChiSq<br>p-value |
| Other | 3.9 (0.0, 9.7) | 2.5 (0.1, 5.0) | 0.6498 | 2.5 (0.0, 5.5) | 1.0 (0.0, 3.0) | 0.4295 |

LHD: Local Health Department; CI: Confidence Interval; PrEP: Preexposure prophylaxis; NACCHO: National Association of County and City Health Officials 2015/2020 Survey
<sup>1</sup> For this analysis, the 2020 survey question "Education and outreach to priority populations for PrEP" is compared to the 2015 survey question "Develop educational materials about PrEP." However, respondents selecting this variable in 2020 may also overlap with respondents who selected the 2015 survey question "Communications activities related to PrEP."
<sup>2</sup> For this analysis, the 2020 survey question "Provide PrEP adherence support" is compared with its closest match in the 2015 survey, "Develop and deliver HIV risk-reduction counseling and behavioral interventions for persons taking PrEP." The authors note that the 2020 question focusing on adherence is more limited in scope than the 2015 question and may represent a smaller number of respondents than those who would choose in 2020 to prioritize funding for the full scope of activities listed in the 2015 question.
<sup>3</sup> For this analysis, the 2020 survey question "Monitoring PrEP uptake" is most closely matched to the 2015 question "Evaluation activities for PrEP-related activities." The authors note that the 2020 question focusing on uptake monitoring may not represent the full scope of evaluation activities covered by the 2015 variable and may represent a smaller number of respondents than those who would choose in 2020 to prioritize funding for the full scope of activities listed in the 2015 question.

### Impact of COVID-19 on LHD Engagement on HIV Testing and PrEP Implementation

At the time of this survey, LHDs were engaged in the COVID-19 pandemic response for over nine months, with significant impact on other essential public health functions performed year-round. LHDs who reported engagement in PrEP implementation (n=158) were asked about the impact of COVID-19 on their PrEP programs (**Table 5**). The most frequently reported impact for all engaged LHDs were a reduced number of staff working on PrEP implementation due to COVID-19 reassignments (41%). One-quarter (24%) of these LHDs (n=65) reported that over 75% of staff were reduced or reassigned from PrEP to work on COVID-19 related activities. Other impacts of COVID-19 on PrEP programming included initiating PrEP in fewer persons (35%), clinic closure (23%), reduced adherence services (15%), and decreased frequency of follow-up visits for PrEP patients (15%). About 12% of the LHDs introduced telehealth services for PrEP and most were LHDs in large jurisdictions (Table 5).

**Table 5.** Impact of COVID-19 on local health department PrEP programs, by jurisdiction size, United States, 2020, weighted response proportions.

| Impact Type | Jurisdiction Size |  |  | Rao-Scott ChiSq<br>p-value |
| --- | --- | --- | --- | --- |
|  | Large<br>(n=43)<br>% (95% CI) | Medium<br>(n=160)<br>% (95% CI) | Small<br>(n=151)<br>% (95% CI) |  |
| Initiating PrEP in fewer persons | 44.4 (28.1, 60.8) | 32.0 (21.4, 42.6) | 34.0 (20.4, 47.6) | 0.5119 |
| Have suspended PrEP initiation visits | 8.3 (0.0, 17.4) | 14.7 (6.6, 22.7) | 4.3 (0.0, 10.1) | 0.1243 |
| Have reduced services to support PrEP adherence and/or retention in care | 13.9 (2.4, 25.4) | 17.3 (8.6, 26.1) | 12.8 (3.2, 22.3) | 0.7420 |
| Reduced staff working on PrEP due to COVID-19 reassignments | 52.8 (36.2, 69.3) | 36.0 (25.0, 47.0) | 42.6 (28.6, 56.5) | 0.3058 |
| Clinic closure | 27.8 (13.0, 42.6) | 30.7 (20.5, 40.9) | 10.6 (1.7, 19.6) | 0.0156 |
| Introduced telehealth services for PrEP | 36.1 (20.2, 52.0) | 12.0 (4.5, 19.5) | 2.1 (0.0, 6.3) | <.0001 |
| Decreased frequency of follow-up visits for PrEP patients | 16.7 (4.3, 29.0) | 18.7 (9.8, 27.5) | 10.6 (1.7, 19.6) | 0.4235 |
| Increased duration of prescriptions for PrEP medications | 8.3 (0.0, 17.4) | 10.7 (3.7, 17.6) | 0 |  |
| Other changes in PrEP service delivery | 5.6 (0.0, 13.0) | 6.7 (0.9, 12.4) | 0 |  |
| Other changes | 25.0 (11.0, 39.0) | 9.3 (2.6, 16.0) | 2.1 (0.0, 6.3) | 0.0039 |
| No impact on PrEP program or activities | 11.1 (0.7, 21.5) | 24.0 (14.2, 33.8) | 34.0 (20.4, 47.7) | 0.0661 |

CI: Confidence Interval; PrEP: Preexposure prophylaxis

### Impact on HIV testing

Eighty-two percent of LHDs who directly provided HIV testing reported reduced or suspended HIV testing in clinical settings during COVID-19, and 55% reported reduced or suspended testing in off-site settings. When asked about challenges related to HIV testing, the most frequently reported response was concerns about social distancing due to COVID-19 (39%), followed by lack of resources such as funding and staff (32%). Twenty-eight percent reported they did not experience any challenges in providing HIV testing.

Overall, 70% of LHDs engaged in PrEP implementation reported reduced (50%) or suspended (20%) provision of HIV and STI testing of PrEP users due to COVID-19, and 30% reported no impact. Among LHDs which reported reductions in HIV and STI testing of PrEP users during COVID-19, approximately one-third (37%) experienced greater than a 50% reduction in testing. Reported adaptations for HIV and STI testing of PrEP users due to COVID-19 included initiation of curbside testing options (8%) and initiation of provision of HIV (7%) or STI (5%) self-testing or self-specimen collection options. Six percent reported offering COVID-19 testing at PrEP care sites (6%).

## Discussion

Local health departments are key partners in implementing and scaling up PrEP in their communities. Notably, since NACCHO’s initial 2015 assessment of LHD PrEP engagement, LHDs were significantly more involved in prescribing PrEP. While the respondents identified areas for improvement, we observed that LHDs were finding their footing as PrEP providers, funders, navigators, and educators, with 41% of engaged LHDs reporting their current role in PrEP implementation was optimal compared with 16% in 2015. Engaged LHDs reported decreased challenges such as those related to navigation for financial assistance and this administrative practice can be shared and scaled-up among U.S. health departments. The breadth and depth of LHDs’ roles related to PrEP implementation highlight that LHDs are assets not only to their communities but also to broader PrEP implementation efforts. It is crucial that LHDs be included by federal, state, and local decisionmakers in plans to increase PrEP access. Funders should create opportunities for LHDs to leverage their experience with technical assistance and peer learning.

Unfortunately, these findings also underscore concerns about implementing PrEP with limited resources, complicated procedures for PrEP medication financial support, and non-adherence of third-party payers to Affordable Care Act requirements for no patient cost-sharing of medications and ancillary services. Respondents reported that increased and sustained funding for PrEP implementation is necessary to protect capacity to support PrEP use especially in public health emergencies such as the COVID-19 pandemic. Additionally, too few LHDs had no engagement in PrEP implementation despite evidence of an increased awareness of their potential roles in PrEP. These findings suggest that additional resources and staff are needed for these LHDs to engage in PrEP implementation. Even before the COVID-19 pandemic, LHDs had limited staff resources for developing community-driven plans for PrEP implementation. LHDs will continue to encounter barriers to initiate engagement in PrEP implementation without specific, streamlined support for planning activities. Engaged LHDs reported implementation planning, outreach, and PrEP education and promotion as optimal activities for PrEP implementation and non-engaged LHDs could benefit from these activities.

Additional needs identified by this assessment include technical assistance and planning support for PrEP implementation. Engaged LHDs are key sources of PrEP education in their communities and are well-positioned to also provide peer-to-peer support to LHDs in earlier stages of engagement. While engaged and unengaged LHDs share some common priorities related to PrEP implementation – including outreach to priority populations, health care provider education, and referral and navigation to PrEP providers – differing resource needs by engagement status indicate areas where non-engaged LHDs may benefit from peer-to-peer learning led by early adopters. However, this work cannot be completed without resources for LHDs to provide technical assistance and/or contribute to regional or national capacity-building efforts.

The impact the COVID-19 pandemic had on LHD PrEP activities reflect the reality that LHDs were not prepared for a public health emergency that tapped their HIV prevention staffing and funding resources. Prior to COVID-19, limited resources were a challenge to LHD engagement in PrEP for HIV prevention, with one-third of engaged LHDs reporting lack of staff as an implementation challenge even before the onset of the pandemic. However, the results of this assessment illustrate that staff redirection to COVID-19 efforts exacerbated these difficulties, stymying ongoing efforts to end the HIV and STI epidemics. During the pandemic, public health agencies depended on the contact tracing expertise of HIV and STD/STI program staff to conduct COVID-19 contact tracing and to train new staff.(7) In May 2020, 78% of the HIV and STD/STI health department workforce was redeployed to the COVID-19 emergency response; in November 2020, at the time this survey was conducted, over 87% of STI programs were supporting or leading COVID-19 contact tracing efforts in their jurisdiction.(8,9) The repercussions of these service interruptions, combined with decreased demand for services, are reflected in estimates of HIV testing outcomes in CDC-funded health departments during COVID-19. In 2020 health departments reported an over one million (47%) reduction in the number of CDC-funded HIV tests and a 30% reduction in persons newly diagnosed with HIV.(10) These findings underscore that HIV and STI prevention efforts were sidelined during the COVID-19 emergency with a public health infrastructure that disproportionately relied on HIV and STI professionals for an emergency response. The expertise of HIV and STI public health workers is important during a public health emergency but decreased HIV and STI services during the COVID-19 response. This underscores the need for models to ensure preparedness, without compromising HIV and STI activities, such as a dedicated emergency response team or upskilling and task shifting of the existing HIV and STI workforce. An adequately resourced public health workforce can provide community outreach, social network approaches to outreach, health promotion, and client navigation to health insurance, medication assistance programs, and PrEP providers.

### Public Health Implications

Robust funding of public health infrastructure can ensure that LHDs and their partners are equipped to provide PrEP access for all who may benefit from it.

### Limitations

While the present study is a unique source of information on HIV prevention and PrEP engagement at the local level, this study is not without limitations. First, the data were self-reported by LHD staff, and NACCHO did not independently verify the data provided. LHD respondents may have provided incomplete, imperfect, or inconsistent information for various reasons which may include skipping questions due to time constraints, estimating responses to reduce burden, or interpreting undefined questions or response options differently. In addition, while the survey methods used in both the 2015 and 2020 surveys were comparable, the wording of some survey items was modified between the two years. This may have impacted respondents’ interpretations, but modifications were typically minor and deemed likely comparable across years. Differences for items presented in this paper are indicated with disclaimers in the tables. The assessment also had a low response rate, which was likely contributed to by LHD capacity constraints during COVID-19. The low response rate may have introduced bias towards more engaged LHDs, LHDs with fewer PrEP responsibilities, or LHDs with fewer COVID-19-related impacts and therefore more time to respond. Future research would benefit from a longer response window and shorter survey, or a series of multiple shorter surveys about specific subtopics. Lastly, the assessment allows a limited point-in-time analysis of the COVID-19 response from October - November 2020 and does not necessarily reflect subsequent impacts of the COVID-19 pandemic on LHD HIV prevention programs through the time of publishing.

## Data Availability

All relevant data are within the manuscript and its Supporting Information files. Supplemental data will be made available through NACCHO.org, CDC.gov, and/or OpenICPSR upon acceptance for publication.

## Acknowledgements

The authors thank all those who contributed to study conception, survey design, data collection, and data interpretation for this assessment and manuscript, especially the late Dawn K Smith, MD, MS, MPH. Dr. Smith’s origination of this assessment during her tenure at CDC was just one of her vast contributions to the development and implementation of pre-exposure prophylaxis (PrEP) for HIV prevention. Her championing of this work and commitment to health equity leaves a lasting impact on the lives of innumerable people.

## Supporting information

S1 Table. Top five resources selected by local health departments as being most helpful for advancing or initiating PrEP implementation, United States, 2020, weighted response proportions.

